# How do People with Multimorbidity Prioritise Healthcare when Faced with Financial Constraints? A Choice Experiment

**DOI:** 10.1101/2023.04.10.23288383

**Authors:** James Larkin, Louise Foley, Shane Timmons, Tony Hickey, Barbara Clyne, Patricia Harrington, Susan M. Smith

## Abstract

Multimorbidity is associated with increased out-of-pocket healthcare costs, making people with multimorbidity more vulnerable to cost-related non-adherence to recommended treatment. This study aimed to understand how people with multimorbidity would prioritise different healthcare services and chronic conditions when faced with potential budget constraints. A cross-sectional online survey incorporating a choice experiment was conducted in Ireland (December 2021 to March 2022). Participants were adults aged 40 years or over with at least one chronic condition. They were asked how they would prioritise their real-world healthcare utilisation if their monthly personal healthcare budget were reduced by 25%. The survey also included questions about real-life experiences of cost-related non-adherence and financial burden. Among the 962 participants, 64.9% (n=624) had multimorbidity. When presented with the hypothetical budget constraint, participants reduced expenditure on ‘other healthcare (hospital visits, specialist doctors, etc.)’, by the greatest percentage (50.2%), and medicines by the lowest percentage (24.5%). Participants with multimorbidity tended to have a condition they prioritised over others. On average, they reduced expenditure for their top-priority condition by 69% less than would be expected if all conditions were valued equally, compared to a reduction in expenditure of 59% more than expected for their least prioritised condition. Participants were asked how important several areas were when making their decisions (scale of 1 – ‘not important at all’ to 5 – ‘very important’). Independence, symptom control and staying alive were rated as the most important decision factors (median=5). Over one third (34.5%, n=332) of participants reported cost-related non-adherence as they had either not attended a healthcare professional or not paid for medication or both in the last year due to cost. Research and clinical care must take greater consideration of the different ways people with multimorbidity respond to high healthcare costs and the impact this has on treatment adherence and health outcomes.

## 1. Introduction

The presence of two or more chronic conditions in an individual (multimorbidity) is one of the biggest challenges for healthcare systems globally (1). The estimated prevalence of multimorbidity in the general population ranges from 13% to 72% depending on setting and age group studied (2) and has been increasing in recent decades (3–5). People with multimorbidity often experience high out-of-pocket healthcare costs, and this is an issue for individuals and healthcare systems internationally (6–10). These high out-of-pocket costs are exacerbated by the negative correlation between multimorbidity and socioeconomic status (11).

High out-of-pocket healthcare costs can lead people with multimorbidity to employ a range of coping strategies (12, 13). One commonly reported coping strategy is cost-related non-adherence to recommended healthcare (14, 15). Adherence refers to the extent to which an individual’s behaviour aligns with recommendations agreed with their healthcare provider (16). Cost-related non-adherence can be problematic for a person’s health and wellbeing (17). It sometimes involves the prioritisation of certain conditions and/or prioritisation of certain healthcare areas (18).

There is extensive literature examining how and why patients prioritise between medications when cost is an issue (19). The underlying reasons for cost-related medication non-adherence are found to be varied. Some have found that patients prioritise short-term benefits (e.g. symptom relief) over long-term benefits (e.g. reduced risk of mortality) (20), though others have found the reverse (18). In relation to how patients actually prioritise between different conditions, when experiencing financial burden, a 2020 study analysed patterns of cost-related medication non-adherence to assess if there were particular groups of conditions that were prioritised (14). They found that cost-related non-adherence was most likely to occur for medications associated with either mental health disorders or respiratory conditions (14). However, none of the above studies have examined how patients prioritise between different areas of healthcare (e.g. primary and secondary care), and also, all have focussed on medications only.

Therefore, this study aimed to understand how people with multimorbidity choose between different healthcare services and chronic conditions when unable to afford healthcare costs. The study also aimed to understand the real-life experiences of healthcare prioritisation for people with multimorbidity.

## 2. Background: Health Entitlements in Ireland

In Ireland, the entitlements system is mixed and there are publicly funded programmes to increase access to healthcare aimed at those with low incomes and/or high healthcare need. There are three main health entitlement categories in Ireland: ‘basic eligibility’, General Practitioner (GP) Visit Card (partial eligibility) and Medical Card (full eligibility) (21). These three categories are mutually exclusive.

- ‘**Basic eligibility**’ entitles people to access the hospital system though with a range of co-payments. At the time of data collection there were upper-limits on charges for inpatient care (€800 annually), emergency department attendance (€100 per visit) and prescription medicines (€114 per month at end of 2021, €100 per month at start of 2022) (22, 23). Patients must pay out-of-pocket for GP visits, with no upper limit. The average cost of a GP visit in Ireland is €54 (24).

- For the **GP Visit Card**, basic eligibility applies but there is also coverage for GP consultation fees (25). Those with household incomes below a threshold are entitled to a GP visit card, with no threshold for adults aged over 70 years (21, 26). As an example of this threshold, for a single person living alone aged under 66 years, there is a weekly income limit (less tax) of €304 (26).

- The **Medical Card** is a means-tested scheme which provides free primary, community and hospital care, and heavily subsidised prescription medicines (27). The income threshold for adults for the Medical Card is approximately 40% lower than the GP visit card threshold (21).

In 2020, 35% of people in Ireland had a Medical Card, 11% had a GP Visit Card and 54% had basic eligibility (28). Also, approximately 46% of the population have private health insurance (29), which facilitates access to private care but generally does not cover GP care. Others pay the full cost out-of-pocket for private care.

## 3 Methods and Data

The study design was a national cross-sectional online survey conducted in Ireland. The survey included: 1) A choice experiment in which people with chronic conditions, including those with a single condition and those with multimorbidity, were asked to prioritise their previous month’s healthcare utilisation within the context of hypothetical budget constraints, and 2) Survey questions about real-life experiences of cost-related non-adherence and financial burden. The study is reported according to the STROBE guidelines (30).

### 3.1 Participants

The inclusion criteria were: a) aged 40 years or over, b) doctor-diagnosed with one or more chronic conditions, c) access to a functional broadband connection and d) access to a computer, laptop, tablet or smart-phone. The age group of 40 years and over was chosen in order to represent some of the younger population living with chronic conditions and multimorbidity. This criterion was also used to increase the likelihood of identifying people who meet the inclusion criteria, as chronic conditions and multimorbidity are less common in those aged under 40 years (31). People with one condition were included to allow for comparison with those with multimorbidity, in line with previous studies of multimorbidity and out-of-pocket healthcare expenditure (Anindya et al., 2021). People were excluded if they: were pregnant or had been pregnant in the previous three months; or had an insufficient level of English language proficiency required to take part. People who were pregnant were excluded as they were likely to have different patterns of healthcare, associated with their pregnancy.

### 3.2 Sampling

The survey was carried out by an independent survey company (32). The company was tasked with sampling a group representative of the public aged 40 years and over with one or more conditions in terms of age, region, sex, socioeconomic status, and area (urban/rural) using stratified random sampling. The company primarily recruited from their pre-established national online research panel of 27,951 people aged 16 years or over. The survey was conducted remotely, online by participants on their own electronic device. The survey was developed using Askia software. Panel members were sent an initial email (S3 Appendix) briefly describing the survey and providing a link to an online portal to participate.

In relation to potential selection bias when using an online approach, a survey by the Central Statistics Office found that 92% of Irish households have internet access in Ireland (33). However, large proportions of certain groups had not used the internet in the three months prior to the survey, particularly those aged over 75 years and over (56%) and the socioeconomically most disadvantage quintile (16%) (34). To increase the response rate among these groups, supplemental door-to-door recruitment was carried out. This involved the researchers from the survey company conducting random door-to-door sampling in randomly chosen locations in Ireland.

When going door-to-door (details in S4 Appendix), recruiters first requested demographic information (S5 Appendix). Then, if the prospective participant met the inclusion criteria, and were in an under-represented group (S4 Appendix), they were provided with study information and how it could be completed online (S6 Appendix).

Participants were offered €10 to complete the survey. Participants who did not complete the survey within 17 days of initial contact were sent a reminder email. Three more reminders were sent, each seven days after the previous email. Data was collected between December 2021 and March 2022. The survey did not contact new prospective participants between 1^st^-16^th^ January because they may have had altered patterns of healthcare usage during the December holiday period.

If participants exited the survey before completion, this was deemed a non-response and answers were not included in analysis. The survey company noted participants’ IP address and device type to prevent people completing the survey twice. This was temporarily stored by Behaviour and Attitudes until data collection was completed, and was only accessible to named individuals at Behaviour and Attitudes.

### 3.3 Choice experiment

A choice experiment is a survey approach whereby participants are presented with several options and the choices they make are used to identify their preferences (35). This choice experiment involved participants outlining their previous month’s healthcare utilisation and associated out-of-pocket expenditure across four different healthcare areas. Participants were then presented with a constrained budget and were asked to decide which areas of their healthcare they would reduce their spending in. The task made it salient that, when faced with a budget constraint, retaining the same level of spending in one area would require reductions in spending in another.

The experiment was not a standard ‘discrete’ choice experiment. Discrete choice experiments involve presenting participants with multiple scenarios that vary systematically by levels of specified attributes. Their preferred scenarios over multiple rounds of choices are used to estimate the weight they assign to those attributes and the levels within them (36). The distinction here is that trade-offs between different attributes are implicit, whereas our design overcomes this limitation and more closely mimics problems faced in real life.

### 3.4 Survey & Choice Experiment Overview

Overall, there were between 59-107 questions, depending on the number of health conditions a participant had. Average survey completion-time was 19 minutes. All questions required a response. A back button was provided to allow participants to change responses. Question types included a mix of closed and open-ended responses. The closed-ended questions were either Likert scale questions or multiple-choice questions. The full questionnaire can be seen in S1 Appendix.

### 3.5 Health, Demographic and Entitlement Information

Participants were presented with a list of 31 chronic conditions which was based on the list used in a national survey of older adults (37). If a participant detailed a condition under ‘other (please specify)’, this person was included only if they had at least one other chronic condition listed in the survey, as it was not feasible to prepare choice experiment questions for conditions reported under the ‘other’ category. Participants were then asked demographic and healthcare entitlement questions.

### 3.6 Healthcare Utilisation

Participants were asked to approximate their healthcare utilisation and the associated out-of-pocket costs for the previous month, broken down into four categories: GP, prescription medicines, primary care, and ‘other healthcare (e.g. hospital visits, specialist doctors, etc.)’. These four categories were chosen as they represent the majority of out-of-pocket healthcare costs experienced by households in Ireland (38). Participants were also asked to break down their expenditure by condition. ‘Multiple/other conditions’ was included as a condition to allow for healthcare utilisation that applies to multiple conditions or other health issues. Participants were asked to include travel expenses in all out-of-pocket healthcare expenditure estimates.

### 3.7 Healthcare Expenditure under Financial Constraints

Participants were then presented with a hypothetical scenario: they experience ‘a large unexpected expense (e.g. tax bill, house repairs, etc.)’, meaning that there is not enough left to pay for their usual level of healthcare utilisation. Participants were then presented with the healthcare expenditure information set at the levels they previously described. However, their budget was now 25% less than the amount it would take to use all of these services as normal. Twenty-five percent was chosen as it was considered sufficient to force people to make decisions that would significantly affect their healthcare utilisation, while low enough to ensure people were not forced to cease most of their healthcare utilisation. Participants were then asked to reduce elements of their healthcare utilisation by enough to at least align their expenditure with the new restricted budget. Participants were not allowed to reduce expenditure other than healthcare (e.g. food or recreation) as the aim of the task was to shed light on which health conditions and healthcare areas people prioritised. Throughout the experiment, participants were provided with explanations and examples (See S1 Appendix).

After completing the choice experiment, participants were given a list of areas that may have informed their prioritisation decisions, and were asked to rate the degree to which each area informed their prioritisation in terms of importance, using a Likert scale ranging from 1 (not important at all) to 5 (very important). The list included maintaining independence, symptom control, clinician advice, staying alive, workload, and ‘other (please specify)’. These options were based on documented priorities of people with multimorbidity (39, 40). Participants were then asked how likely they would be to use alternative ways of paying for healthcare (e.g. use savings, borrow money) if they were presented with this scenario in their real lives based on a scale of 1 (very unlikely) to 5 (very likely). These were adapted from the options in the composite measure of financial burden for patients with stage three colorectal cancer (41).

The composite measure of financial burden for patients with stage three colorectal cancer was also used to understand participants’ real life experience of healthcare related financial burden (S1 Appendix, Q7-8). The measure was adapted for this purpose. This included a question asking participants how much they worry about financial problems that have resulted from the cost of their healthcare on a scale of 1 (not at all) to 7 (very much).

Participants were then asked about the number of occasions they did not purchase medicines in the previous 12 months because of cost and number of occasions they did not attend a healthcare professional in the previous 12 months because of cost.

### 3.8 Pilot

The survey company and the researchers conducted a pilot study with 60 participants. Based on the pilot responses the following changes were made: minor technical issues were addressed, estimated completion time was revised, the wording of questions 4 and 4b were slightly revised with more detail added to ensure understanding, and an example of healthcare-related travel expenses was added to question 3.1.1. The changes were not considered substantial and therefore the pilot participants were included in the final sample.

### 3.9 Patient & Public Involvement (PPI)

The survey was developed in consultation with a PPI panel of people with multimorbidity. To develop the financial burden scenario, members of the panel were asked to provide examples of scenarios that are likely to cause financial burden to a large proportion of people with multimorbidity. The panel was also presented with the survey in its entirety and asked a series of questions to ensure comprehension (S7 Appendix). One of the members of the PPI panel (co-author, TH) was included as a co-researcher and contributed to conceptualization, methodology, and reviewing and editing.

### 3.10 Data Analysis

A multimorbidity count was developed by combining some of the 31 conditions in the questionnaire to give 21 broader conditions (S2 Appendix), based on data availability and a previous Irish Longitudinal Study of Ageing study on multimorbidity (42). For example, stroke, peripheral vascular disease and transient ischemic attack were combined into vascular disease. A participant’s multimorbidity count was based on this list of 21 broader conditions.

Frequencies and descriptive statistics were used to describe the demographics of the participants and their reported real-life experience of financial burden.Descriptive statistics were used to describe the average monthly healthcare utilisation and healthcare expenditure. Average percentage difference was used to describe difference in healthcare expenditure under financial constraints. These descriptive statistics were analysed based on type of healthcare (e.g. prescription medicines, general practice etc.) and number of chronic conditions (one condition, two conditions and three or more conditions). The categories ‘two conditions’ and ‘three or more conditions’ were included to align with previous studies of multimorbidity and out-of-pocket healthcare expenditure (7).

Out-of-pocket healthcare expenditure is likely to be positively skewed. This was accounted for by using percentage reduction as the primary outcome variable, as the percentage reduction for large expenditures is likely to be similar to the percentage reduction for small expenditures due to the fixed goal of a 25% reduction in overall out-of-pocket healthcare expenditure. Also, if there are a large number of respondents with no expenditure in certain healthcare areas, this will not affect the mean percentage reduction.

To assess prioritisation between conditions, proportionate unit reduction was used. This method was adopted as no standard analysis for this type of data was identified in the literature. Only those with expenditure on two or more conditions (including multiple/other conditions) were included. In the presented analysis of proportionate unit reduction for named conditions, a condition was only included if at least 100 participants with expenditure on two or more conditions had expenditure on that condition. This was in order for there to be sufficient numbers to facilitate meaningful analysis. Proportionate unit reduction was determined by calculating the percentage reduction for each condition and summing those percentages. Then, this total percentage was divided by the number of conditions (including *multiple/other conditions* as a condition). A proportionate unit reduction of a value less than -1 (e.g. -1.5) represents a deprioritisation of the respective condition and a proportionate unit reduction of a value greater than -1 (e.g. -0.4) represents a prioritisation of the respective condition. An example of the proportionate unit reduction method for a participant can be seen in S8 Appendix.

Proportionate unit reduction was also used to assess whether participants generally treat conditions equally or tend to prioritise a condition. To assess this, the average proportionate unit reduction for all participants’ most prioritised condition and least prioritised condition, was taken. The specific condition that was prioritised or de- prioritised varied from participant to participant. ‘Multiple/other conditions’ was not included when averaging the proportionate unit reduction for most and least prioritised conditions. This was because prioritising this category may mean that an individual is prioritising their overall health and not just ‘other conditions’ specifically.

Medians and interquartile ranges were used to analyse Likert scale questions. A conventional content analysis (43) of open ended questions 4b, 5b and 7 was conducted. These were free-text questions related to what informed participants’ prioritisation decisions, alternatives to sacrificing healthcare and participants’ real-life experiences of making sacrifices due to healthcare costs. The first stage involved (JL) reading and re-reading the responses. The second stage involved creating codes under which responses could be categorised. The third stage consisted of assigning data to the codes. A second author (LF) crosschecked this process independently. Discrepancies were addressed through discussion.

### 3.11 Outlier Management

All healthcare expenditure data that was greater than two standard deviations from the mean was reviewed by a health services researcher (JL) and a GP (SMS). Expenditure that JL and SMS considered unlikely to be feasible was removed. Reasons for this included allocation of excessively high costs for a particular area that is known not to incur such costs. This led to expenditure data for 10 of the 962 participants being removed. An example of an outlier that was removed was €555 for three GP visits. The same process was applied for cost-related non-adherence data.

For cost-related non-adherence, responses that were removed were more than nine standard deviations greater than the mean. This led to data for five of the 962 participants being removed (more details in S9 Appendix).

### 3.12 Ethics Statement

Ethical approval for this study was obtained from the RCSI University of Medicine and Health Sciences Research Ethics Committee (reference number: 202104018). The data was fully anonymised before being accessed by the research team.

Participants provided written consent for their responses to be used for research purposes.

## 4 Results

### 4.1 Participant Characteristics

In total, 962 respondents completed the survey. Of the 962 who completed the survey, 13.0% (n=125) were recruited face-to-face to complete the online survey (further details of face-to-face recruits and online recruits can be seen in S4 Appendix). A flow chart of the response rate can be seen in Fig 1.

**Fig 1.**
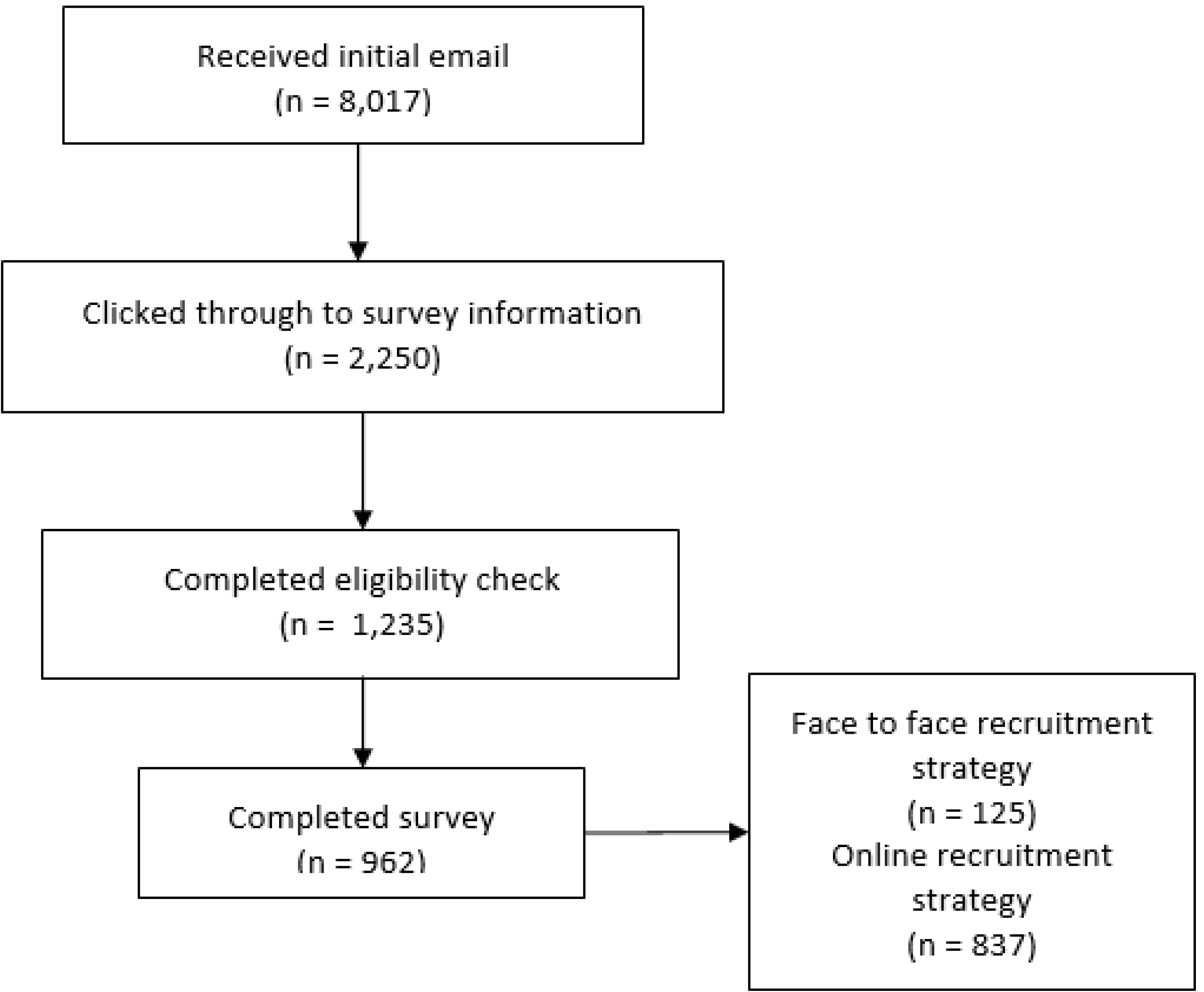
Flow chart of response rate

Overall, 54.8% (n=527) of participants were female, 16.2% (n=156) had a primary school education or less, 55.1% (n=530) were aged between 40-59 years old, and 64.9% (n=624) had multimorbidity (2+ conditions). Table 1 provides details of the sample characteristics. The sample was relatively representative when compared with those aged 50 years and over with one or more condition in the 2016 sample of the Irish National longitudinal Study on Ageing (S10 and S11 Tables).

**Table 1.** Demographic and entitlement characteristics of sample

|  | Overall (n=962)<br>% (n) | One<br>condition<br>(n=338)<br>% (n) | Multimorbidity |  |
| --- | --- | --- | --- | --- |
|  |  |  | Two<br>conditions<br>(n=265)<br>% (n) | Three or more<br>conditions<br>(n=359)<br>% (n) |
| Age (years) |  |  |  |  |
| 40-49 | 22.8 (219) | 31.6 (107) | 18.9 (50) | 17.3 (62) |
| 50-59 | 32.3 (311) | 32.8 (111) | 38.1 (101) | 27.6 (99) |
| 60-69 | 29.8 (287) | 26.0 (88) | 30.2 (80) | 33.1 (119) |
| 70-79 | 11.8 (113) | 8.3 (28) | 11.3 (30) | 15.3 (55) |
| 80-89 | 2.5 (24) | 1.2 (4) | 1.1 (3) | 4.7 (17) |
| 90+ | 0.8 (8) | 0.0 (0) | 0.4 (1) | 1.9 (7) |
| Sex |  |  |  |  |
| Female | 54.8 (527) | 48.5 (164) | 56.2 (149) | 59.6 (214) |
| Male | 45.1 (434) | 51.4 (174) | 43.4 (115) | 40.3 (145) |
| Other | 0.1 (1) | 0.0 (0) | 0.4 (1) | 0.0 (0) |
| Education |  |  |  |  |
| Primary/none | 16.2 (156) | 13.0 (44) | 15.2 (40) | 20.1 (72) |
| Secondary | 36.7 (353) | 36.1 (122) | 37.5 (99) | 36.8 (132) |
| Third/higher | 47.0 (452) | 50.9 (172) | 47.3 (125) | 43.2 (155) |
| Location |  |  |  |  |
| Urban (5000+ people) | 53.1 (511) | 58.6 (198) | 50.2 (133) | 50.1 (180) |
| Rural (<5000 people) | 46.9 (451) | 41.4 (140) | 49.8 (132) | 49.9 (179) |
| Private Health Insurance |  |  |  |  |
| Yes | 54.7 (526) | 55.6 (188) | 54.0 (143) | 54.3 (195) |
| No | 45.3 (436) | 44.4 (150) | 46.0 (122) | 45.7 (164) |
| Healthcare Entitlements |  |  |  |  |
| Medical Card | 42.3 (407) | 34.3 (116) | 41.9 (111) | 50.1 (180) |
| GP Visit Card | 10.8 (104) | 8.6 (29) | 10.2 (27) | 13.4 (48) |
| Neither | 46.9 (451) | 57.1 (193) | 47.9 (127) | 36.5 (131) |

### 4.2 Current Expenditure and Response to Hypothetical Budget Constraints: Healthcare Areas

Participants’ average monthly healthcare expenditure was highest for ‘other healthcare (hospital visits, specialist doctors, etc.)’ at €47.15 (SD=321.54) and lowest for primary care (physiotherapy, occupational therapist, psychologist) at €13.50 (SD=58.09). When participants were presented with the hypothetical budget constraints, they chose to reduce other healthcare expenditure, by the greatest percentage (50.2%), and medicines by the lowest percentage (24.5%). When looked at by number of conditions, this pattern was the same for all groups – ‘other healthcare’ was reduced by the greatest percentage and medicines by the least. The one exception was those with one condition, who reduced primary care by the least. Further details broken down by number of conditions are in Table 2. Medians and interquartile ranges for the same variables are provided in S8 Table. Number of participants who reached the payment limits for medicines (described in section 2.2) is provided in S9 Table. Overall, 6.2% (N=43) of those who had expenditure on medicines reached a relevant medicine payment threshold associated with their entitlements, for example the upper limit of €114 per month on medications (more details in section 2).

**Table 2.** Expenditure reductions in response to financial constraints

|  |  | Previous month's healthcare expenditure<br>Mean (SD) | Monthly healthcare expenditure after choices made under financial constraints<br>Mean (SD) | Mean percentage reduction in expenditure |
| --- | --- | --- | --- | --- |
| Overall | GP | €37.21 (71.10) | €22.96 (48.11) | 38.3% |
|  | Medicines | €33.67 (51.17) | €25.43 (40.20) | 24.5% |
|  | Primary Care (physio, occupational therapist, psychologist) | €13.50 (58.09) | €7.34 (33.86) | 45.6% |
|  | 'Other Healthcare (hospital visits, specialist doctors, etc.)' | €47.15 (321.54) | €23.47 (203.47) | 50.2% |
| One Condition | GP | €21.84 (38.40) | €14.94 (27.56) | 31.6% |
|  | Medicines | €19.59 (31.39) | €14.36 (25.90) | 26.7% |
|  | Primary Care | €5.43 (29.05) | €4.26 (23.27) | 21.5% |
|  | 'Other Healthcare' | €23.04 (88.36) | €10.66 (52.49) | 53.7% |
| Two Conditions | GP | €45.44 (79.50) | €27.68 (54.43) | 39.1% |
|  | Medicines | €34.88 (43.34) | €27.30 (36.06) | 21.7% |
|  | Primary Care | €16.47 (66.52) | €6.65 (28.51) | 59.6% |
|  | 'Other Healthcare' | €26.45 (87.49) | €12.12 (45.62) | 54.2% |
| Three or more Conditions | GP | €45.61 (84.79) | €27.01 (56.80) | 40.8% |
|  | Medicines | €46.05 (66.20) | €34.49 (50.51) | 25.1% |
|  | Primary Care | €18.92 (70.08) | €10.74 (44.18) | 43.2% |
|  | 'Other Healthcare' | €85.14 (512.36) | €43.92 (326.26) | 48.4% |

### 4.3 Response to Hypothetical Budget Constraints: Chronic Conditions

Overall, 40.7% (n=392) of participants had expenditure on two or more conditions (including *multiple/other conditions* as one condition). In general, when participants were choosing which condition to reduce expenditure on, they did not reduce expenditure equally across conditions. The most and least prioritised conditions varied between individuals but when they are considered in general terms and multiple/other conditions is excluded, the average proportionate unit reduction for the most prioritised condition as a measure across all participants, was -0.31 (SD=0.51), which represents a reduction that was 69% less than expected if all conditions were treated equally. The average proportionate unit reduction for the least prioritised condition was -1.59 (SD=1.03), which represents a reduction 59% more than expected if all conditions were treated equally.

For conditions that were reported by at least 100 participants (Table 3), the most prioritised condition was high blood pressure or hypertension (n=162), which had a proportionate unit reduction of -0.75 (SD=0.87) and a mean absolute percentage reduction of 38.0%. The least prioritised condition was multiple/other conditions (n=234), which had a proportionate unit reduction of -1.46 (SD=1.19) and a mean absolute percentage reduction of 58.0%. The full list of conditions and their respective reductions can be seen in S3 and S4 Tables.

**Table 3.** Proportionate unit reduction of expenditure for each condition with more than 100 participants

| Chronic condition | Average proportionate unit reduction (SD) | Mean percentage reduction in expenditure |
| --- | --- | --- |
| High blood pressure or hypertension (N=162) | -0.75 (0.87) | 38.0% |
| High cholesterol (N=137) | -0.83 (0.89) | 43.5% |
| Any emotional, nervous or psychiatric problems, such as depression or anxiety (N=102) | -0.84 (0.96) | 33.0% |
| Arthritis (including osteoarthritis, or rheumatism) (N=106) | -1.21 (1.04) | 66.1% |
| Multiple/other conditions (N=234) | -1.46 (1.19) | 58.0% |

### 4.4 Influences on Prioritisation Choices

Participants rated the importance of different factors when reducing their healthcare expenditure, using a scale ranging from 1 (not important at all) to 5 (very important). Staying alive, maintaining independence, and symptom control were the most important factors for participants, each with a median rating of 5. Doctors’ advice and treatment burden both had a median rating of 4. A more detailed breakdown can be seen in S5 Table. Participants were asked if there was ‘anything else’ that informed their prioritisation decisions and provided with a free-text box. Additional factors were reported by 15.6% (n=150) of respondents. Of these, 7.2% (n=69) of participants reported that the availability of an alternative form of care/therapy would inform their decision and 2.6% (n=25) said they would consider the intensity of the illness at the given point in time. Full details of alternative considerations are in S6 Table.

### 4.5 Alternative Cost-Saving Choices & Real-Life Healthcare Related Financial Burden

Participants were asked, if they were presented with this scenario in their real-life (not having enough money to access one’s usual level of healthcare), how likely did they think there were to sacrifice parts of their usual healthcare usage on a scale of 1 (very unlikely) to 5 (very likely). The median response was 3 (IQR=2-4) with 46.1% of respondents stating that they were either likely or very likely to sacrifice parts of their usual healthcare usage. People were more likely to use savings, cut down on recreational spending, or cut down on general non-healthcare expenses to pay for their healthcare, rather than sacrifice parts of their usual healthcare usage (S1 Table). These were also the strategies that people were more likely to have previously used in their own lives; 46.3% (n=445) of participants had cut down on general expenses to pay for healthcare. Overall, 60.7% (n=584) of participants had had to use a non-healthcare cost-saving measures at some stage due to healthcare cost challenges. Further details of likelihood of using hypothetical cost saving decisions are in Fig 2 and further details of real-life cost saving decisions are in Fig 3. The distribution of responses for hypothetical cost saving decisions are in S1 Table.

**Fig 2.**
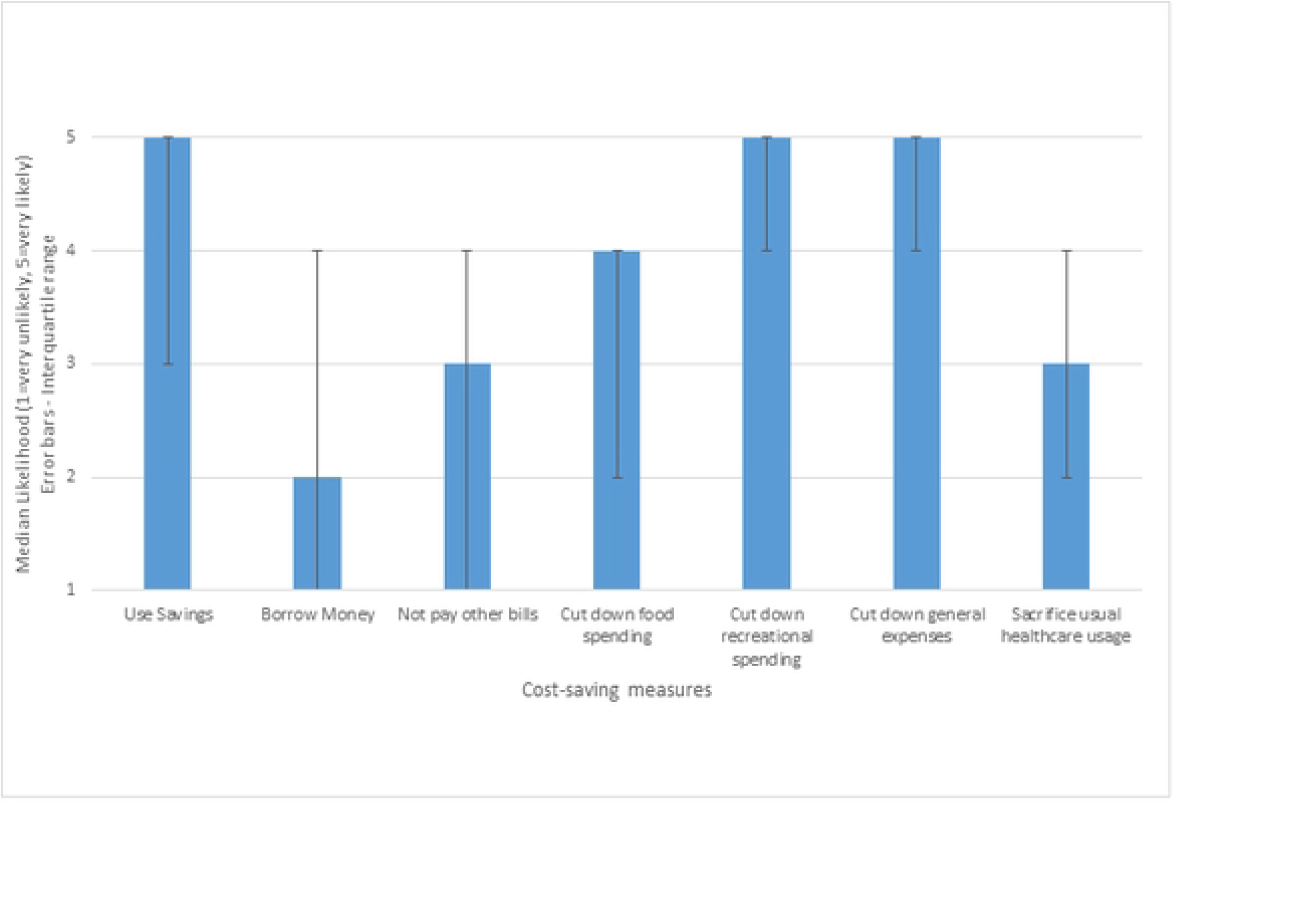
Hypothetical likelihood of cost saving measures when faced with healthcare cost challenges

**Fig 3.**
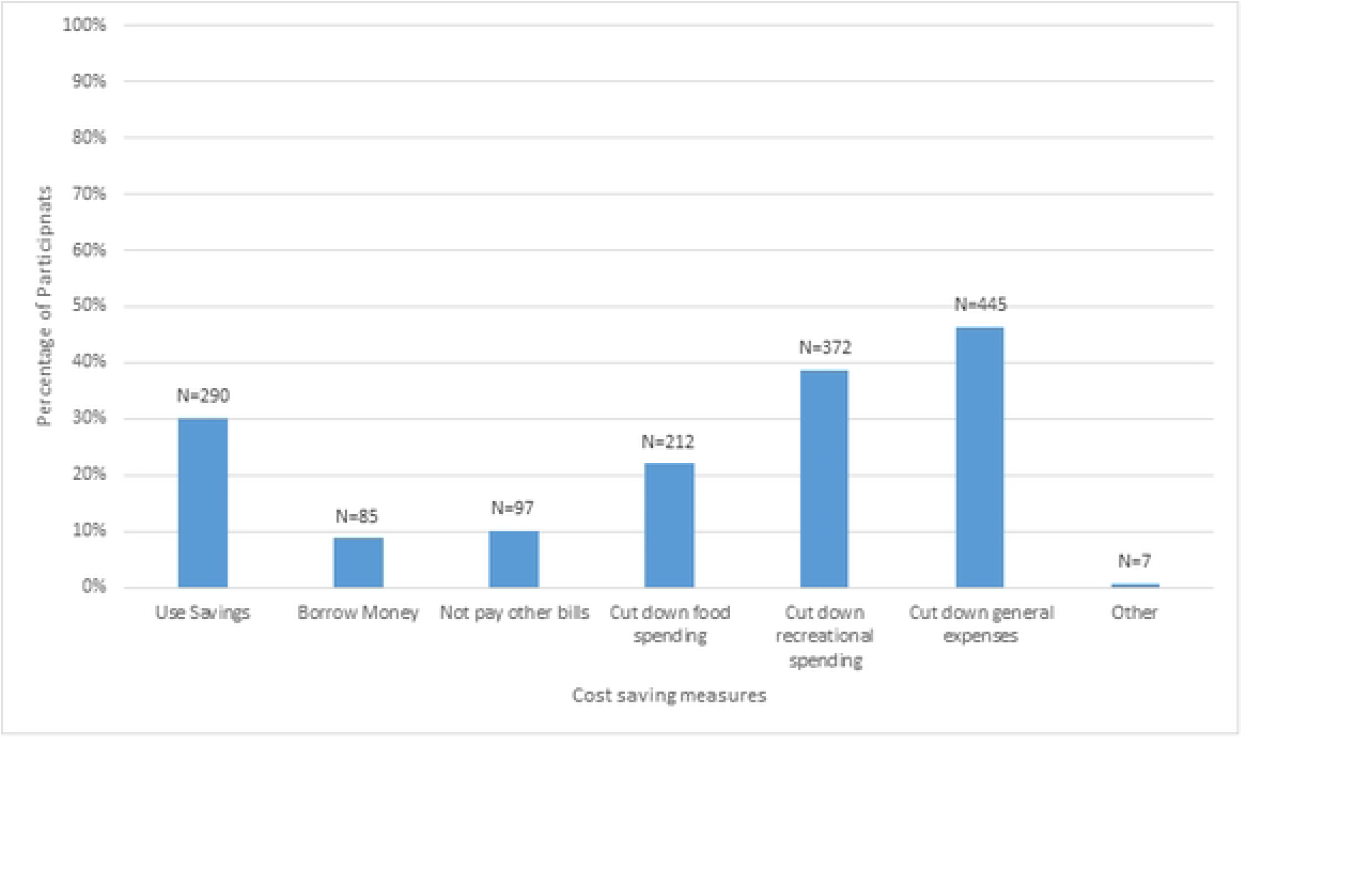
Real-life experiences of cost saving measures when faced with healthcare cost challenges

Participants were also asked if there were ‘other’ alternatives they would have used to allow them to protect their usual level of healthcare, and provided with a free-text box. For 15.9% (n=153) of participants, there were other alternatives they would use. Of these, 4.7% (n=45) of participants said they would increase their income (e.g. sell assets or work more), 3.4% (n=33) said they would change their behaviour to attempt to improve their health (e.g. through exercise, diet etc.) and avoid the need for healthcare, 0.7% (n=7) said they would try to negotiate with their healthcare provider, and 0.4% (N=4) said they would access care in another country. Full details of ‘other’ alternatives to reducing healthcare expenditure are in S7 Table.

Participants were asked how much they worry about financial problems that have resulted from the cost of their currently recommended healthcare treatments on a scale of 1 (not at all) to 7 (very much). The median response was 4 (IQR=2-6). The distribution of responses can be seen in S2 Table.

### 4.6 Cost-Related Non-Adherence

For healthcare visits, 31.1% (N=299) of participants reported not attending a healthcare professional in the previous 12 months because of costs, and among those that did not attend, the average number of occasions that this occurred in the previous 12 months was 2.4 (SD=1.8). For cost-related non-adherence to medications, 15.5% (N=149) of participants reported not buying a medication in the previous 12 months because of cost, and among those that did not buy a medication, the average number of occasions that this occurred in the previous 12 months was 2.7 (SD=2.9). Overall, 34.5% (N=332) either reported not attending a healthcare professional or not buying a medication in the previous 12 months or both because of cost. Table 4 provides more details of cost-related non-adherence broken down by number of conditions and medical card status.

**Table 4.** Cost-related non-adherence/attendance in previous 12 months

|  | Ever not attended<br>healthcare<br>professional in<br>previous 12 months<br>due to cost<br>% (N) | No.<br>occasions<br>Mean (SD) | Ever not bought<br>medicine in<br>previous 12<br>months due to<br>cost<br>% (N) | No.<br>occasions<br>Mean<br>(SD) |
| --- | --- | --- | --- | --- |
| Holders of medical<br>card or GP visit card |  |  |  |  |
| Overall | 27.0 (138) | 2.4 (2.0) | 15.7 (80) | 2.8 (3.4) |
| One Condition | 20.7 (30) | 2.3 (2.6) | 12.4 (18) | 2.7 (5.5) |
| Two Conditions | 25.4 (35) | 2.1 (1.7) | 15.9 (22) | 2.5 (2.1) |
| Three+ Conditions | 32.0 (73) | 2.6 (1.8) | 17.5 (40) | 3.0 (2.8) |
| Holders of neither<br>card |  |  |  |  |
| Overall | 35.7 (161) | 2.4 (1.7) | 15.3 (69) | 2.6 (2.2) |
| One Condition | 31.6 (61) | 2.2 (1.3) | 14.5 (28) | 2.1 (1.4) |
| Two Conditions | 40.2 (51) | 2.5 (1.7) | 15.7 (20) | 3.6 (3.3) |
| Three+ Conditions | 37.4 (49) | 2.3 (2.0) | 16.0 (21) | 2.1 (1.3) |
| All Participants |  |  |  |  |
| Overall | 31.1 (299) | 2.4 (1.8) | 15.5 (149) | 2.7 (2.9) |
| One Condition | 26.9 (91) | 2.6 (1.8) | 13.6 (46) | 2.4 (3.5) |
| Two Conditions | 32.5 (86) | 2.4 (1.7) | 15.8 (42) | 3.0 (2.8) |
| Three+ Conditions | 34.0 (122) | 2.5 (1.9) | 17.0 (61) | 2.7 (2.4) |

## 5. Discussion

This study presents a systematic evaluation of how people with chronic conditions, including multimorbidity, make trade-offs between their chronic conditions and between different healthcare services when experiencing financial constraints. This study also reports on wider cost-related non-adherence to recommended treatments for people living with chronic conditions; over a third of participants reporting that in the previous 12 months they had either not attended appointments or not accessed medicines due to costs. In the choice experiment, the area of healthcare that participants prioritised the most was medicines, while they prioritised ‘other healthcare (hospital visits, specialist doctors, etc.)’ the least. Participants reported that they would reduce their expenditure for ‘other healthcare’ by over twice as much as for medicines. For those with multimorbidity, people tended to have a condition they demonstrated was of greatest importance to them, reducing their expenditure for that condition by, on average, less than a fifth of what was expected if they had reduced expenditure across all conditions proportionately. For their least prioritised condition, they reduced their expenditure by more than double what was expected. The areas that mainly informed participants’ decision making when prioritising services were ‘staying alive’, ‘symptom control’ and ‘maintaining independence’.

The finding that people would prioritise their medicines over GP visits, primary care and ‘other healthcare’ usage is consistent with their reports of their actual experience of cost-related non-adherence. While 16% of people had reported previous non-adherence to medicines due to cost, almost twice as many people (31%) reported previous non-attendance at a healthcare professional for the same reason.

Nonetheless, in the wider context, healthcare was a high priority for participants. Participants stated they were more likely to use savings, cut down on recreational spending, or cut down on general expenses than reduce their healthcare expenditure. This highlights a risk for those with chronic conditions and multimorbidity who have lower incomes and higher out-of-pocket healthcare expenditure (6) and therefore may not have the capacity to use these mechanisms. Participants reported being less likely to borrow money to address the hypothetical financial constraints than reduce their healthcare utilisation, which may mean that there is a threshold up to which participants protect their healthcare spending if they can. Free text responses provided interesting examples of other alternatives to reducing healthcare expenditure considered by a minority of participants, such as negotiating with the healthcare provider, accessing care in another country, or increasing income through more paid work or selling personal items. When participants were asked about their real-life use of alternatives, the responses mirrored their hypothetical likelihood: people were most likely to report that they had used savings, cut down on recreational spending and/or cut down on general expenses.

### 5.1 Comparison with Existing Literature

Our findings that participants prioritised medication expenditure aligns with the findings of a 2017 study of older adults in Ireland with polypharmacy (prescribed five or more medications), which reported that 96% believed strongly in the necessity of their medication (44). A Swedish study of ‘frail elderly patients in primary care’ also found that the vast majority believed in the necessity of their medication (45). “Doctors’ advice” was important for the majority of participants in the choice experiment when deciding which area of healthcare to prioritise. The 2017 study also found that people who were prescribed more medications (which is correlated with higher number of conditions) had stronger beliefs in the necessity of their medications (44), which aligns with our findings that people with higher number of conditions were less likely to reduce their expenditure on medicines compared with other healthcare areas. It has also been reported that as functional health status decreases, belief in the necessity of medicines prescribed for multimorbidity increases, further highlighting the perceived centrality of medicines in the management of multiple chronic conditions (46). However, given that there is very little research examining the perceived necessity of, or beliefs about, other healthcare areas, it is difficult to put these prioritisation decisions into a wider context. Previous research that looked at medication non-adherence found that mental health disorders are among the most vulnerable group of conditions to cost- related non-adherence (14). Contrary to that, this study found that ‘Any emotional, nervous or psychiatric problems, such as depression or anxiety’ was one of the most prioritized areas and therefore least likely to be subject to cost-related non- adherence. Nonetheless, this study aligns with several qualitative studies showing that people prioritise individual conditions when engaging in medication taking behaviours (47, 48).

Despite the finding that medicine was the area that participants prioritised in response to hypothetical budget constraints, about one in six participants had engaged in cost-related medication non-adherence in the previous year. Some studies have found that a small increase in costs can lead some people to reducing their medicine use, but for other people, they continue to adhere to medicines even when experiencing significant financial burden (19). There are mixed results in the literature on whether independence, symptom control or reduced mortality risk are more important to patients when prioritising medicines (19, 49). This study shows that all of these areas are reported to be very important for people when prioritising their conditions and their healthcare. This finding is somewhat expected given the clinical diversity that is inherent in multimorbidity, and the potential for people to be living with multiple co-occurring conditions which simultaneously increase their risk of mortality (50) and present symptoms which reduce quality of life and independence (51).

The proportion of participants (16%) who reported cost-related medication non-adherence in the previous 12 months is generally higher than rates reported internationally. A study of cost-related prescription medication non-adherence in the previous 12 months among older adults in 11 developed countries found rates of between <3% (France, Norway, Sweden, Switzerland and the UK) and 17% (USA) (52). However, the setting and populations differed between the studies with the international study including all adults aged 55 years and older (52). It also included prescription medicines only, while this study looked at ‘medication you needed for your treatment.’

### 5.2 Strengths & Limitations

Previous studies have explored the priorities of individuals with multimorbidity using qualitative interview methods to explore the lived of experience of people with multimorbidity (49, 53), whereas the current study involved a choice experiment and a large national sample targeting people with multimorbidity. The set-up of the choice experiment in this study is a strength as it facilitates decision-making that closely replicates real world decision-making. It does this by identifying the participant’s actual healthcare utilisation and then constraining participants to choose between several alternatives at once. Also the questions were adapted from previous studies and informed by a multimorbidity PPI panel. Nonetheless, hypothetical bias is a limitation of this study.

The sample was another strength, as it was representative when compared with those aged 50 years and over with one or more condition in the Irish National longitudinal Study on Ageing that were identified in a 2016 study of adults aged 50 years or over (8). This was achieved through the additional door-to-door recruitment to access groups who may be less represented in survey panels. The high rate of non-completion (22.1%) amongst those who started the survey, is a limitation and is likely a result of the length of the survey (54, 55).

In Ireland, some areas of healthcare (e.g. medicines and hospital stay) have a threshold, over which the state pays the excess. Participants at these thresholds may have been less likely to reduce expenditure in the respective area, as it may have involved making a very significant change to their healthcare access. However, only a small number of participants met these thresholds. Given that the study was conducted in Ireland, and the complexity of decision-making relating to adherence and financial burden, generalisability of these types of studies is somewhat limited without local adaptations and considerations. A further limitation related to responses to the Likert scale questions about participant reasons for decision making in the prioritisation exercise. These questions demonstrated a ceiling effect (56). Use of an alternative scale or ranking process may have provided more informative findings.

### 5.3 Policy and Practice

This study shows the distinct healthcare areas and conditions people prioritise when they are asked to address financial constraints. That, coupled with the high levels of reported real-life cost-related non-adherence, suggests a need for policymakers to consider the impact of financial burden on treatment choices and adherence in development of clinical guidelines and policy. Currently NICE guidance on the management of multimorbidity recommends addressing patient priorities in general terms but does not give specific guidance on the management of multimorbidity when people may have to make healthcare choices when experiencing financial burden (57). Our study’s findings show that people with multimorbidity prioritise certain services and conditions and de-prioritise others. NICE and other clinical guidance for multimorbidity could incorporate specific advice to healthcare workers to consider financial burden when discussing patient priorities.

Healthcare workers could also be advised to explore explicitly how costs might affect patients’ adherence to recommended treatments. This could be conducted in clinical practice as part of a cost-of-care conversation between patients and healthcare workers, which would involve a discussion of financial costs and/or coverage related to health or healthcare (58). These cost-of-care conversations should be promoted, as they rarely take place despite having the potential to reduce patient costs (59) and increase adherence (60), thereby increasing the clinical benefit of prescribed treatments. Consideration of referral to a welfare rights advisor or social worker could also be considered when people are experiencing financial burden (61). To prevent financial burden, given that user fees are strongly associated with a reduction in healthcare utilisation (62), reducing the payment barriers to accessing care can address these high rates of cost-related non-adherence and non-attendance at a system level.

### 5.4 Conclusion

This study’s findings suggest that many people with chronic conditions including multimorbidity would choose to prioritise their medicines when experiencing financial burden. They also tend to have a condition that they have demonstrated is of greatest importance to them. Participants reported a high prevalence of real-life cost-related non-adherence to treatment. Clinical guidelines should incorporate guidance in this area, advising healthcare workers to consider discussing how patients might prioritise between conditions or between healthcare areas, as well as how costs might affect their decision-making. Overall, research and clinical care must take greater consideration of the potentially harmful effects of high healthcare costs experienced by people with multimorbidity, which can lead to healthcare choices that can have negative long-term effects on health.

## Data Availability

Researchers interested in using the data associated with this study may access the data for free from the following sites: Irish Social Science Data Archive (ISSDA) at University College Dublin https://www.ucd.ie/issda/data/choiceexperiment/

## Acknowledgements

We are thankful to the Health Research Board Ireland [CDA- 2018-003] for funding this research. We are grateful to the public and patient representatives who provided constructive feedback on several aspects of this study. We would like to acknowledge Dr. Féidhlim McGowan and Prof. Eamon O’Shea for advice on the design of the choice experiment. We are also grateful to Behaviour and Attitudes Limited for carrying out the survey.

